# A quantitative assessment of *Staphylococcus aureus* community carriage in Yuma, Arizona

**DOI:** 10.1101/2022.05.24.22275524

**Authors:** Benjamin Russakoff, Colin Wood, Monica R. Lininger, Steven D. Barger, Robert T. Trotter, Sara Maltinsky, Mimi Mbegbu, Briana Coyne, David Panisello Yague, Shari Kyman, Kara Tucker-Morgan, Kathya Ceniceros, Cristina Padilla, Kevin Hurtado, Ashley Menard, Francisco Villa, Heidi A. Wayment, Crystal Hepp, Tara Furstenau, Viacheslav Fofanov, Cindy M. Liu, Talima Pearson

## Abstract

*Staphylococcus aureus* is a frequent cause of mild and severe infections that occur when these commensal bacteria penetrate the outer layers of skin or mucosa. As most *S. aureus* infections are the result of autoinfection, and community-acquired infections are increasingly common, it is important to better understand *S. aureus* colonization characteristics in the community setting. Using standard culture technique and a quantitative PCR assay (SaQuant), we detected and quantified *S. aureus* across the nares, throat, and palm of 548 community-dwelling individuals in southwestern Arizona. Using culture-based methods, we detected *S. aureus* colonization in the nares of 26.3% of individuals (n = 144); however, the combination of two detection methods across multiple body sites resulted in much higher prevalence than has been reported previously. Overall, 65.9% of participants were colonized, with significantly higher prevalence in males (compared to females) and non-Hispanics (compared to Hispanics), with this pattern especially evident in nares and throat samples. Colonizing quantities in the nares were slightly higher in males and significantly greater among non-Hispanics. The clear sex and ethnicity patterns warrant further investigation in order to identify and leverage protective factors that may drive these disparities. In the nares, *S. aureus* density was the highest, most variable, and correlates with colonization in other body sites such as throat and palm. Our results demonstrate that screening by culture-based methods only can miss individuals colonized by *S. aureus* and that previous carriage statistics are likely underestimates. By including a highly sensitive quantitative assay, this work provides a roadmap towards more comprehensive and accurate characterization of *S. aureus* carriage and the potential for more effective mitigation.

**AUTHOR SUMMARY:** Effective disease control and prevention is tied to pathogen identification and understanding reservoirs. *Staphylococcus aureus* infection prevention efforts and protocols are based upon decades of research on colonization patterns and associated links to subsequent infection. Unfortunately, efforts to prevent *S. aureus* infections have been met with diminishing returns, suggesting significant gaps in fundamental knowledge of colonization. However, this knowledge and resulting protocols, are founded upon culture-based detection. By employing a new quantitative PCR assay on samples from three body sites in 548 individuals, we can characterize colonization more comprehensively than previous studies by describing both prevalence and pathogen quantity. Our highly sensitive detection resulted in an overall prevalence of 65.9%. Higher quantities were associated with the nares and were highest among non-Hispanic males (86.9%). Overall prevalence was much higher than has been previously documented. Common research practices, such as culture-based detection from a single body site, may misclassify over half of colonized persons. Future studies incorporating quantitative data, especially with longitudinal sampling at more body sites will provide a more wholistic understanding of community carriage, colonization dynamics, and likelihood of autoinfection and transmission.

## INTRODUCTION

*Staphylococcus aureus* is a human commensal and opportunistic pathogen that causes skin and soft-tissue infection, bone and joint infection, bacterial endocarditis, and bacteremia (Williams 1963; Tong et al. 2015). Methicillin-resistant *S. aureus* (MRSA) has received particular attention due to treatment challenges surrounding antibiotic resistance and its high burden on the health-care system, though recent data suggest that invasive methicillin-susceptible *S. aureus* (MSSA) cases significantly outnumber MRSA cases (Centers for Disease Control and Prevention 2020; Koeck et al. 2019; Jackson et al. 2020; Crandall et al. 2020). Increased effort to control life-threatening *S. aureus* infections in hospitals has resulted in a steady decline in cases over the past 15 years (Kourtis et al. 2019). However, progress in the health-care setting is out of step with increasing cases of community-onset bacteremia (Kourtis et al. 2019; Asgeirsson, Thalme, and Weiland 2017). These patterns suggest an increasingly relevant role of the community as a reservoir for *S. aureus* transmission and persistence.

Self-infection is the most common cause of *S. aureus* infection (von Eiff et al. 2001; Wertheim et al. 2005). Carriage is thus not only a marker for prevalence in the community, but also an important risk factor for infection. Estimates of *S. aureus* carriage vary widely with geographic location, sex, age, and ethnicity. In the United States, carriage is estimated at approximately one-third of the population (Gorwitz et al. 2008; Graham, Lin, and Larson 2006; Kuehnert et al. 2006; Gualandi et al. 2018; Curry et al. 2016). Carriage is higher in children than in adults, peaking between the ages of six and twelve. Throughout adulthood, carriage is mostly stable, declining slightly with age (Gorwitz et al. 2008; Graham, Lin, and Larson 2006; Erikstrup et al. 2019; Holtfreter et al. 2016; den Heijer et al. 2013). Males are more likely to be colonized than females (den Heijer et al. 2013; Holtfreter et al. 2016; Erikstrup et al. 2019; Curry et al. 2016; Gorwitz et al. 2008; VandenBergh et al. 1999; Kuehnert et al. 2006; Mainous et al. 2006). Ethnicity-based trends are less clear-cut but suggest that Hispanic and non-Hispanic whites have a similar prevalence that is higher than that of non-Hispanic blacks (Mainous et al. 2006; Kuehnert et al. 2006; Gorwitz et al. 2008).

*S. aureus* colonizes numerous body sites, the most common of which are the nares, hand, perineum, and throat (Wertheim et al. 2005). The nares are commonly reported as the most frequent site of colonization and are thought to provide a reservoir for transfer to other sites (Williams 1963; Hogan et al. 2020; J. Kluytmans, van Belkum, and Verbrugh 1997; Rohr et al. 2004). Indeed, decolonization of the nares has been shown to decrease colonization at other body sites (Reagan et al. 1991). Nasal carriage is also an important risk factor for infection during surgery (J. A. J. W. Kluytmans et al. 1995; Kalmeijer et al. 2000). Although the importance of nasal carriage is well-documented, an increasing number of studies have examined throat and nasal carriage in parallel and report similar (Young et al. 2017) or greater prevalence in the throat (Erikstrup et al. 2019; Sollid et al. 2014; Nilsson and Ripa 2006; Hamdan-Partida, Sainz-Espuñes, and Bustos-Martínez 2010), suggesting that the throat may play an under-appreciated role as a body site reservoir.

Given that the quantity of *S. aureus* among colonized persons has implications for autoinfection and transmission, current binary *S. aureus* carriage characterization likely provide an incomplete representation of *S. aureus* burden. However, quantitative aspects of carriage at a given body site have not been widely studied. For example, the quantity of *S. aureus* in the nares is positively associated with postsurgical infection risk (Calia et al. 1969; J. A. J. W. Kluytmans et al. 1995; White 1961a), with higher likelihood of transmission to household members (Ehrenkranz 1964), and with greater dissemination of *S. aureus* into the environment (White 1961b). It has also been observed that, relative to males, nasal carriage in women is characterized by smaller *S. aureus* quantities and a higher likelihood of a false-negative carriage classification using conventional culturing methods. Thus, bacterial quantity is an important yet understudied feature of colonization, and combining independent quantitative and culture-based assessments of *S. aureus* should improve the validity of investigations of possible sex-based disparities in nasal carriage and putative sex-based determinants of *S. aureus* transmission (Liu et al. 2015). Although most quantitative studies for *S. aureus* have been culture-based, this approach does not lend itself well to quantification due to resource intensity and limited throughput.

To better understand binary and quantitative community-based carriage, we apply a quantitative PCR assay to samples collected from a large cohort of community members in Yuma, Arizona (Barger et al. 2020; Pearson et al. 2019; 2021). Based on data collected during the validation of a new highly sensitive and specific qPCR assay for the accurate detection and quantification of *S. aureus* (Wood et al. 2021), we expected to achieve higher detection sensitivity compared to culture-based detection. We further hypothesized, in line with existing literature, that the nares would be the most frequently colonized body site, with the highest colonization loads; that prevalence and quantity would be greater in males compared to females; that prevalence and quantity between Hispanics and non-Hispanics would be similar; and finally, that prevalence and quantity would decrease slightly with age.

## METHODS

### Study Overview

This study is part of a larger investigation aimed at understanding carriage and transmission within social groups in a population on the US/Mexico border (Pearson et al. 2021; Barger et al. 2020; Pearson et al. 2019). Study staff recruited participants in naturally occurring social groups of two or more persons and supervised the self-collection of swabs from three body sites: the nares, throat, and palm. Participants additionally filled out a questionnaire which, among other variables, documented their demographic information. Any person older than six months was eligible to participate and received a gift card as an incentive. Adult participants provided informed consent and children provided assent. Responses for younger children were completed by an adult proxy.

Participants were recruited in naturally occurring groups of 2 or more persons and included individuals ranging from infancy to adults as old as 93 years. Data were collected from 565 participants from March of 2019 through March of 2020. For the analyses within this manuscript, 17 participants were excluded for incomplete data on sex, age, ethnicity, or assay, leaving a complete sample of 548 participants. From these participants, 608 of the samples from the three body sites were positive for *S. aureus* using culture and molecular-based assessments.

### Sample collection

Two double-tipped BBLCultureSwab swabs were used to sample each of the three body sites (each body site was therefore sampled twice). Study staff demonstrated and explained the swabbing protocol to each participant before providing them with swabs and assisted participants when necessary. Study staff performed the swabbing for young children or elderly participants who could not or chose not to self-collect. When sampling the nares, participants were instructed to insert the double tip swab into the anterior nares and rotate it as they moved it around, sampling each nostril for ten seconds with the same swab. When sampling the throat, participants were instructed to swab the back of their tongue, mouth, and cheeks for twenty seconds while rotating the swab. When sampling the palm, participants were instructed to rub the swab over the palm of their hand and between their fingers for twenty seconds. During sampling, study staff counted aloud and reminded participants to rotate the swab and cover the entire body site during the sampling of each body site to maintain sample collection consistency across individuals.

### Sample storage, transportation, and culturing

After collection, all swabs were stored and transported on ice to the laboratory. In the laboratory, one of the two swabs from each body site was used for culturing. The selected swab was stored for no longer than 24 hours at 4°C before culturing to maximize the likelihood of cell survival (Panisello Yagüe et al. 2021). Each such swab was streaked onto CHROMagar *S. aureus* media and incubated for 24 hours at 37°C. Plates were read after incubation to assess for *S. aureus* growth. Colonies that appeared in the pink to mauve color range were considered to be *S. aureus*. One colony from each swab was isolated and sequenced for species verification and all suspected *S. aureus* colonies were stored at -70°C in 20% glycerol. The second swab from each body site, which did not undergo culturing, was stored at -70°C until direct DNA extraction.

### DNA extraction

DNA was isolated from suspected positive *S. aureus* colonies using the Qiagen DNEasy Blood and Tissue kit with modified lysis for gram positive bacteria. Each sample was streaked for isolation and a lawn from a single colony was grown for extraction. The remaining swab samples (one from each body site) that were not used for culturing were extracted on a Thermofisher KingFisher Flex instrument using an Applied Biosystems MagMAX™ DNA Multi-Sample Ultra 2.0 Kit with a final elution volume of 50μL.

### qPCR assay

The SaQuant qPCR assay was run on 1μL aliquots of DNA extract, using an Applied Biosystems QuantStudio 7 and 12 real-time PCR machines following the published protocol (Wood et al. 2021). Standard curves for quantification were constructed using a minimum of five 10-fold serial dilutions, and at least one standard curve was included in each qPCR run. A quantification value for each sample was generated from its associated standard curve(s) using the QuantStudio software. Data are presented as genome equivalents per reaction.

### Data analyses

Statistical analyses and data visualization was carried out using R version 1.3.1093 (R Core Team 2018). Violin plots, pie charts, and Venn diagrams were generated using the data visualization package ggplot2 version 3.3.3 (Wickham 2016). Mann-Whitney tests were conducted to determine significant differences between *S. aureus quantities* across dichotomous independent variables (culture detection, sex, ethnicity). Because of correlated data, the Friedman test was used to detect differences in quantity between the three body sites (nares, throat, and palm), with post-hoc Nemenyi pairwise comparisons. Two-proportion z-tests were used to determine statistically significant differences between prevalence among sexes (males and females) and ethnicities (Non-Hispanic and Hispanic). Chi-squared tests were utilized to determine statistically significant differences in prevalence across age classes, while the Kruskal-Wallis rank sum test with post-hoc Mann-Whitney tests, if warranted, were used to compare quantities across age classes. Linear regression models were generated to determine potential associations between colonization and *S. aureus* quantities at one body site with another. Tests yielding a two-tailed p-value ≤ 0.05 were considered statistically significant.

## RESULTS

### Study participants

The 548 participants had an average age of 32.1 years. There were 96 participants under the age of 18. Most participants (50.5%) were married, a slight majority (54.9%) were female, and most (72.8%) reported Hispanic ethnicity. Sociodemographic characteristics of participants are included in Table 1.

**Table 1.**
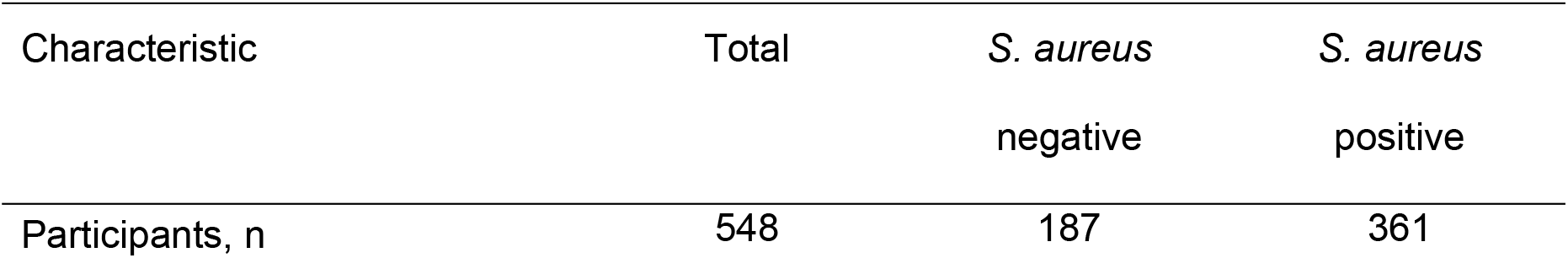
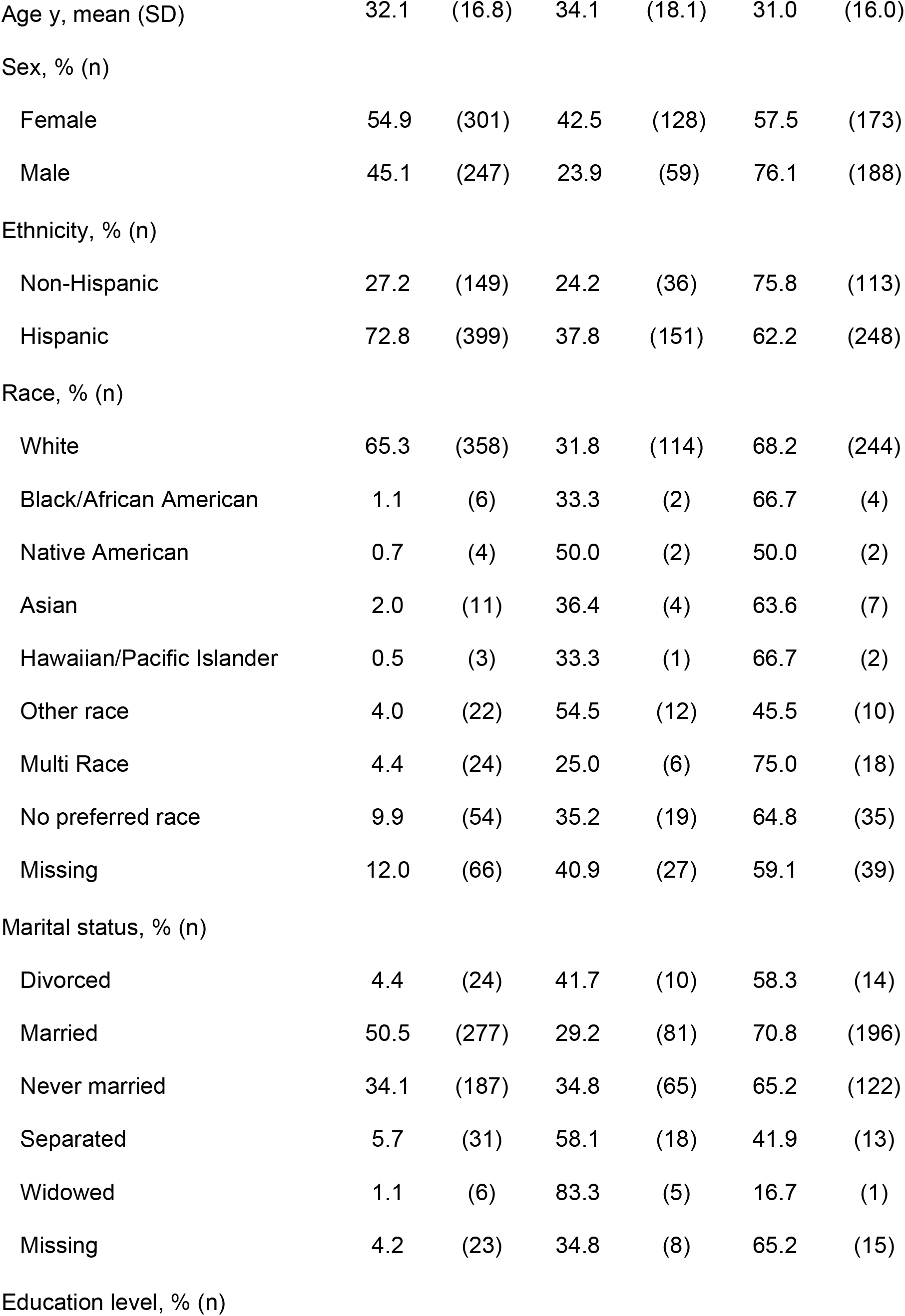
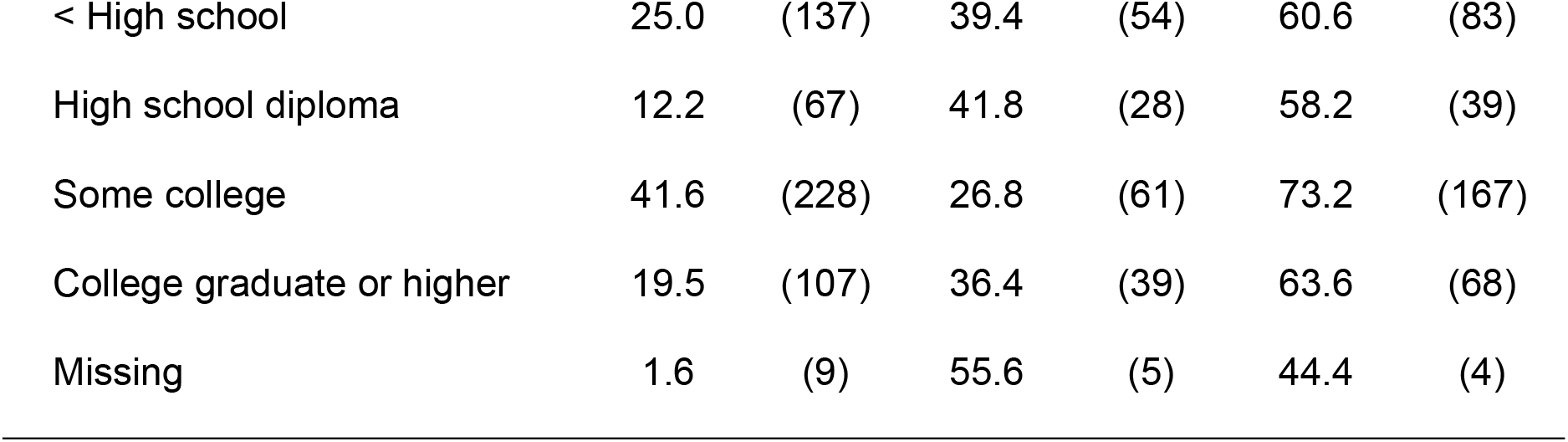
Sample Baseline Characteristics.

| Characteristic | Total | *S. aureus*<br>negative | *S. aureus*<br>positive |
| --- | --- | --- | --- |
| Participants, n | 548 | 187 | 361 |

|  |  |  |  |  |  |  |
| --- | --- | --- | --- | --- | --- | --- |
| Age y, mean (SD) | 32.1 | (16.8) | 34.1 | (18.1) | 31.0 | (16.0) |
| Sex, % (n) |  |  |  |  |  |  |
| Female | 54.9 | (301) | 42.5 | (128) | 57.5 | (173) |
| Male | 45.1 | (247) | 23.9 | (59) | 76.1 | (188) |
| Ethnicity, % (n) |  |  |  |  |  |  |
| Non-Hispanic | 27.2 | (149) | 24.2 | (36) | 75.8 | (113) |
| Hispanic | 72.8 | (399) | 37.8 | (151) | 62.2 | (248) |
| Race, % (n) |  |  |  |  |  |  |
| White | 65.3 | (358) | 31.8 | (114) | 68.2 | (244) |
| Black/African American | 1.1 | (6) | 33.3 | (2) | 66.7 | (4) |
| Native American | 0.7 | (4) | 50.0 | (2) | 50.0 | (2) |
| Asian | 2.0 | (11) | 36.4 | (4) | 63.6 | (7) |
| Hawaiian/Pacific Islander | 0.5 | (3) | 33.3 | (1) | 66.7 | (2) |
| Other race | 4.0 | (22) | 54.5 | (12) | 45.5 | (10) |
| Multi Race | 4.4 | (24) | 25.0 | (6) | 75.0 | (18) |
| No preferred race | 9.9 | (54) | 35.2 | (19) | 64.8 | (35) |
| Missing | 12.0 | (66) | 40.9 | (27) | 59.1 | (39) |
| Marital status, % (n) |  |  |  |  |  |  |
| Divorced | 4.4 | (24) | 41.7 | (10) | 58.3 | (14) |
| Married | 50.5 | (277) | 29.2 | (81) | 70.8 | (196) |
| Never married | 34.1 | (187) | 34.8 | (65) | 65.2 | (122) |
| Separated | 5.7 | (31) | 58.1 | (18) | 41.9 | (13) |
| Widowed | 1.1 | (6) | 83.3 | (5) | 16.7 | (1) |
| Missing | 4.2 | (23) | 34.8 | (8) | 65.2 | (15) |
| Education level, % (n) |  |  |  |  |  |  |
| < High school | 25.0 | (137) | 39.4 | (54) | 60.6 | (83) |
| High school diploma | 12.2 | (67) | 41.8 | (28) | 58.2 | (39) |
| Some college | 41.6 | (228) | 26.8 | (61) | 73.2 | (167) |
| College graduate or higher | 19.5 | (107) | 36.4 | (39) | 63.6 | (68) |
| Missing | 1.6 | (9) | 55.6 | (5) | 44.4 | (4) |

### Comparison of *S. aureus* detection methods

For each body site, we compared culture and qPCR for detection of *S. aureus*. Culturing was not specific for detecting *S. aureus*, resulting in false positives that were excluded from this sample set after whole genome sequence comparisons. Culturing was also not as sensitive as SaQuant, resulting in 297 out of 608 positive samples (48.8%) detected via culture compared to 553/608 (91.0%) detected by SaQuant (Fig 1A). The SaQuant assay is new with impressive sensitivity and specificity values calculated by running the assay against 533 *S. aureus* isolates and 10 non-*aureus Staphylococcus* isolates as well as *in-silico* against 1,818 *S. aureus* genomes and 1,834 non-aureus *Staphylococcus* genomes (Wood et al. 2021). However, as this assay had not been assessed outside the initial validation study (Wood et al.), we further evaluated the sensitivity and specificity of the assay with these data.

**Fig 1.**
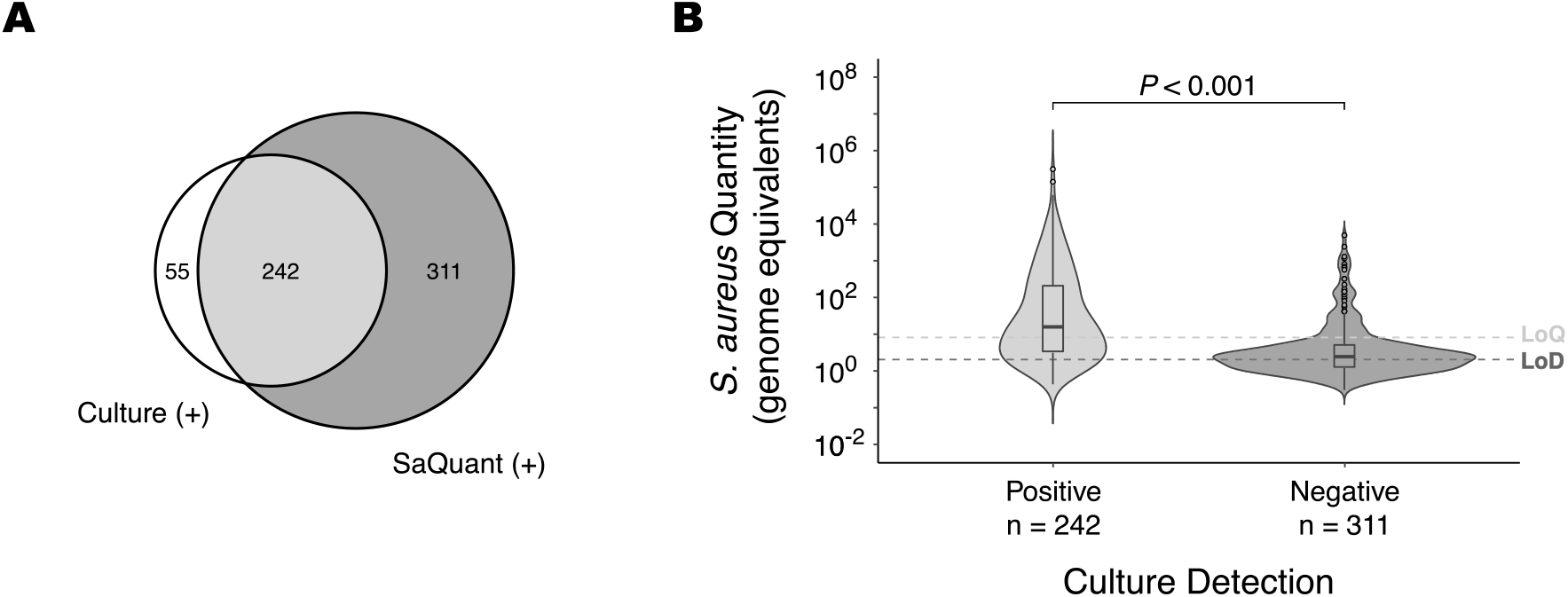
Comparison of culture and SaQuant for *S. aureus* detection. A) Proportion of samples (n=608) that were positive with only culture, both culture and SaQuant qPCR, and only SaQuant qPCR. A large proportion of samples were exclusively detected by SaQuant while only a few were exclusively picked up by culture. B) Distribution of *S. aureus* quantity in 553 samples positive for the SaQuant assay shows higher quantities in samples that were culture positive compared to culture negative samples. Detection and quantitative comparisons suggest that the SaQuant qPCR assay is more reliable for *S. aureus* detection due to a higher likelihood of detecting low quantities. Differences in *S. aureus* quantities between culture positive and negative samples were statistically significant according to the Mann-Whitney test (*P* < 0.001). *S. aureus* amounts below the limit of detection (LOD) of between 3 and 5 genomic copies can still be detected, albeit at less than a 95% confidence level. Likewise, amounts below the limit of quantification (LOQ) of 8.27 genomic equivalents will be quantified with less accuracy.

Across the three body sites, there were 55 culture positive but PCR negative samples. As only a single replicate of each sample was tested against SaQuant, we expected samples with low *S. aureus* quantities to be stochastically detected, leading to a small proportion of false negatives. As we had culture and whole genome sequences from the paired swab for these 55 samples, we were able to confirm a 100% sequence match with the primer and probe target sites to show that genomic mutations are not responsible for the lack of amplification. Low quantities of *S. aureus* on a sample swab are also likely to account for the 311 samples that were culture negative, but detectable via qPCR as they had a significantly lower quantity of *S. aureus* (Fig 1B).

For assurance that SaQuant was not producing false positive results, we performed amplicon sequencing on samples with the highest amounts of *S. aureus* DNA as detected by the SaQuant assay. We targeted 23 *S. aureus* specific amplicons that did not amplify in other *Staphylococcus* species (using *in silico* PCR). The targets were successfully amplified in each of the samples and BLASTn (Zhang et al. 2000) searches of the sequences resulted in only hits to *S. aureus*.

### *S. aureus* at different body sites

Culture and qPCR detection were used to determine *S. aureus* presence at each sample site (Fig 2A). The SaQuant assay revealed a higher *S. aureus* prevalence than culture, suggesting higher sensitivity. However, culture was able to detect *S. aureus* in some samples that were negative for SaQuant, as well as a subset of samples that did not undergo qPCR testing due to insufficient DNA reserves. Therefore, combining both methods provided the highest level of detection. Overall detection of *S. aureus* was similar at the nares and throat body sites, with respective presence in 233 and 234 of 548 (42.5 and 42.7%) participants compared to only 151/548 (27.6%) for palm samples. Body site results from Fig 2A were used to determine prevalence on a participant basis (Fig 2B). Individuals positive at any site were considered positive for *S. aureus*. The SaQuant assay was more sensitive than culture at all body sites and combining both methods across three body sites resulted in *S. aureus* prevalence of 65.9% (361/548) of participants. Overall prevalence represents a broad overview of *S. aureus* colonization but does not account for the prospect of colonization at multiple sites. Hence, prevalence results were dissected to indicate the proportion of individuals colonized exclusively at one body site, two sites, and all three sites (Fig 2c). Consistent with Fig 2A and 2b, a combination of detection methods yielded the highest prevalence of *S. aureus* colonization. Individuals colonized exclusively at a single body site accounted for 49.9% of positive participants (180/361), while 29.1% of positive participants were colonized at two body sites (105/361) and 21.1% (76/361) were colonized at all three sites. A major benefit of a qPCR assay is the ability to quantify the target of interest present in a sample. In this study, we utilized the SaQuant assay to quantify the *S. aureus* load for sample swabs collected from the nares, throat, and palm (Fig 3). *S. aureus* quantity was significantly different across all three body sites (Friedman test, *P* < 0.001). Nares samples exhibited higher quantities compared to both throat (*P* < 0.001) and palm samples (*P* < 0.001), while throat samples had higher quantities compared to palm samples (*P* < 0.001).

**Fig 2.**
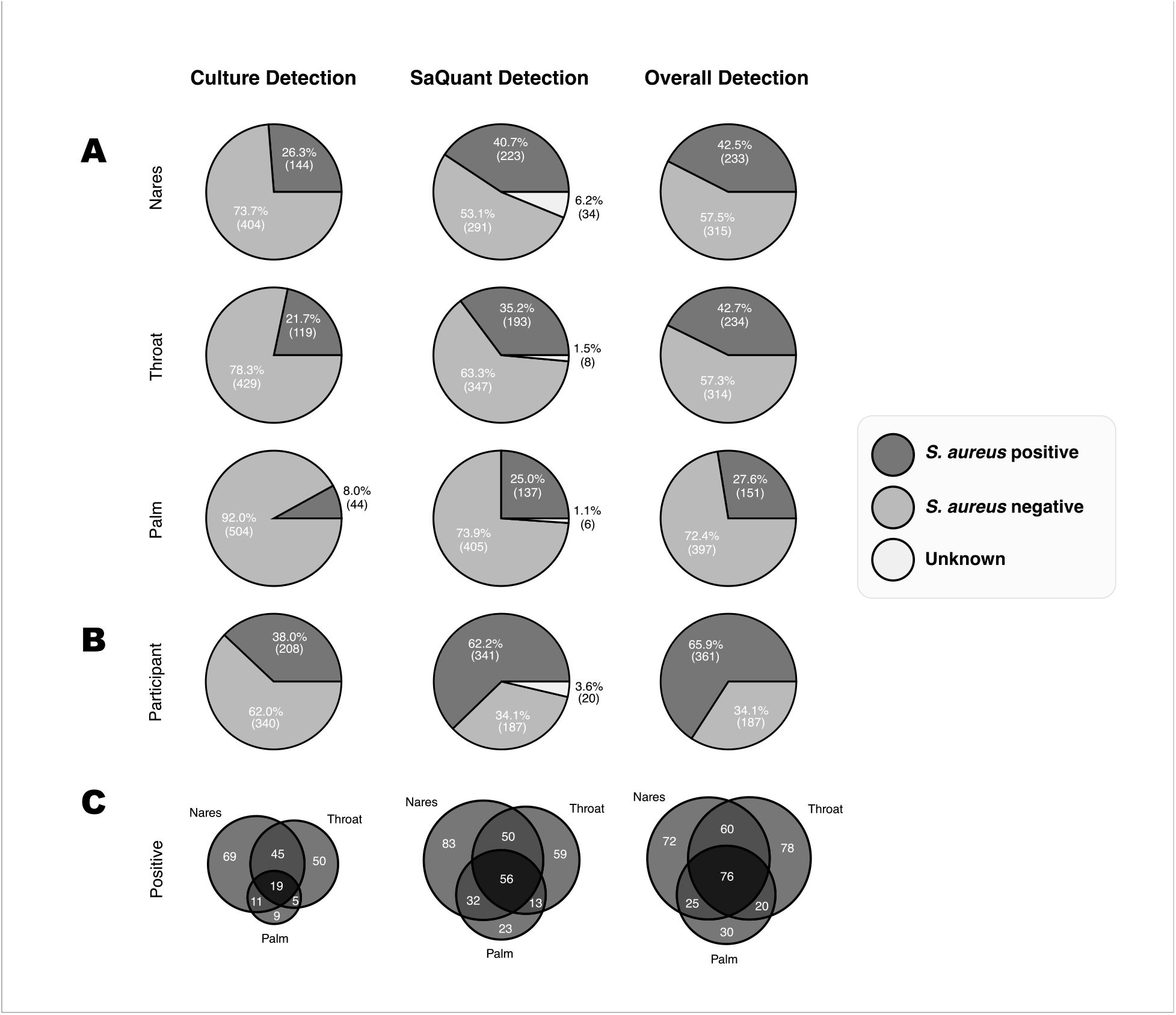
Prevalence across sites for 548 participants. A) Consistent with Fig 1, SaQuant shows greater sensitivity in detecting *S. aureus* at each body site. Combining culture and SaQuant results provides the highest detection sensitivity. A subset of samples could not be evaluated with SaQuant, as DNA extraction failed. B) When data are combined across all 3 body sites, detection with SaQuant is greater than for culture, but combining SaQuant with culture provides the greatest overall detection. For culture, combining data across 3 sites results in a prevalence estimation that is slightly greater than most published population accounts. When detection methods and body sites are combined, the overall prevalence is 65.9%. C) The number of individuals with carriage at single or multiple body sites. Individual samples that did not have a pair at the other body sites were excluded.

**Fig 3.**
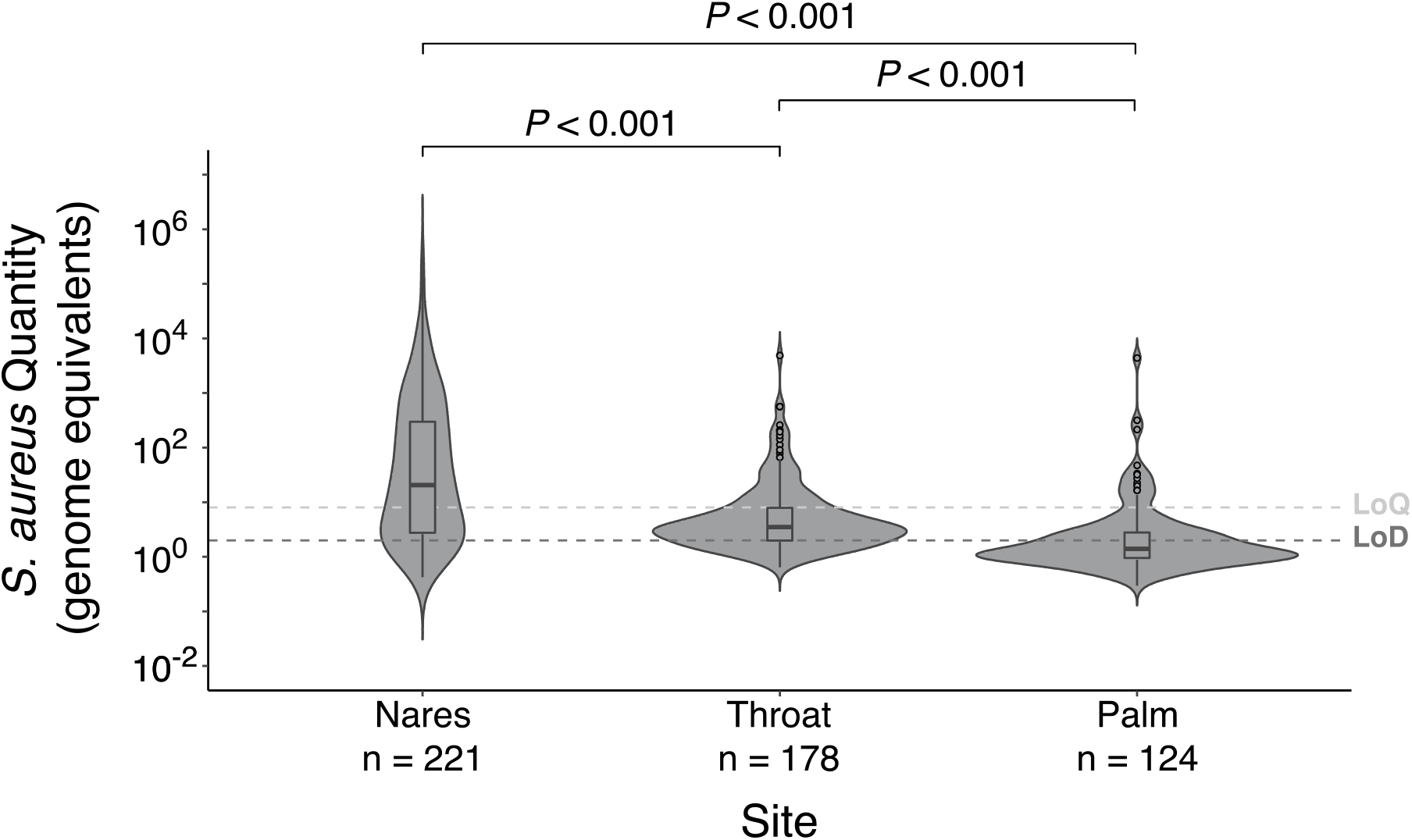
Quantities at different body sites. Quantities differ at each body site (Friedman test, *P* < 0.001). Specifically, the quantity in nares is greater than the throat and palm (Nemenyi test, *P* < 0.001) and throat quantities are greater compared to the palm (Nemenyi test, *P* < 0.001).

### *S. aureus* colonization and sex, ethnicity, and age

We evaluated sex and ethnicity-based disparities in colonization and quantity. Sex-based disparities were of particular interest given differences in male and female bacterial colonization. As previously discussed, *S. aureus* prevalence was highest when qPCR and culture results were combined. Taking this combined approach, males showed significantly higher *S. aureus* prevalence compared to females at all individual body sites, as well as overall colonization prevalence (i.e., any positive *S. aureus* result; Fig 4A). Differences in *S. aureus* quantity between males and females at the nares (*P* = 0.094), throat (*P* = 0.695), and palm (*P* = 0.152) were not statistically significant (Fig 4B). The apparent discrepancy with overall site-based prevalence is likely due to the small sample size for quantitative analysis, as some samples were exclusively detected by culture and therefore have no associated quantity value.

**Fig 4.**
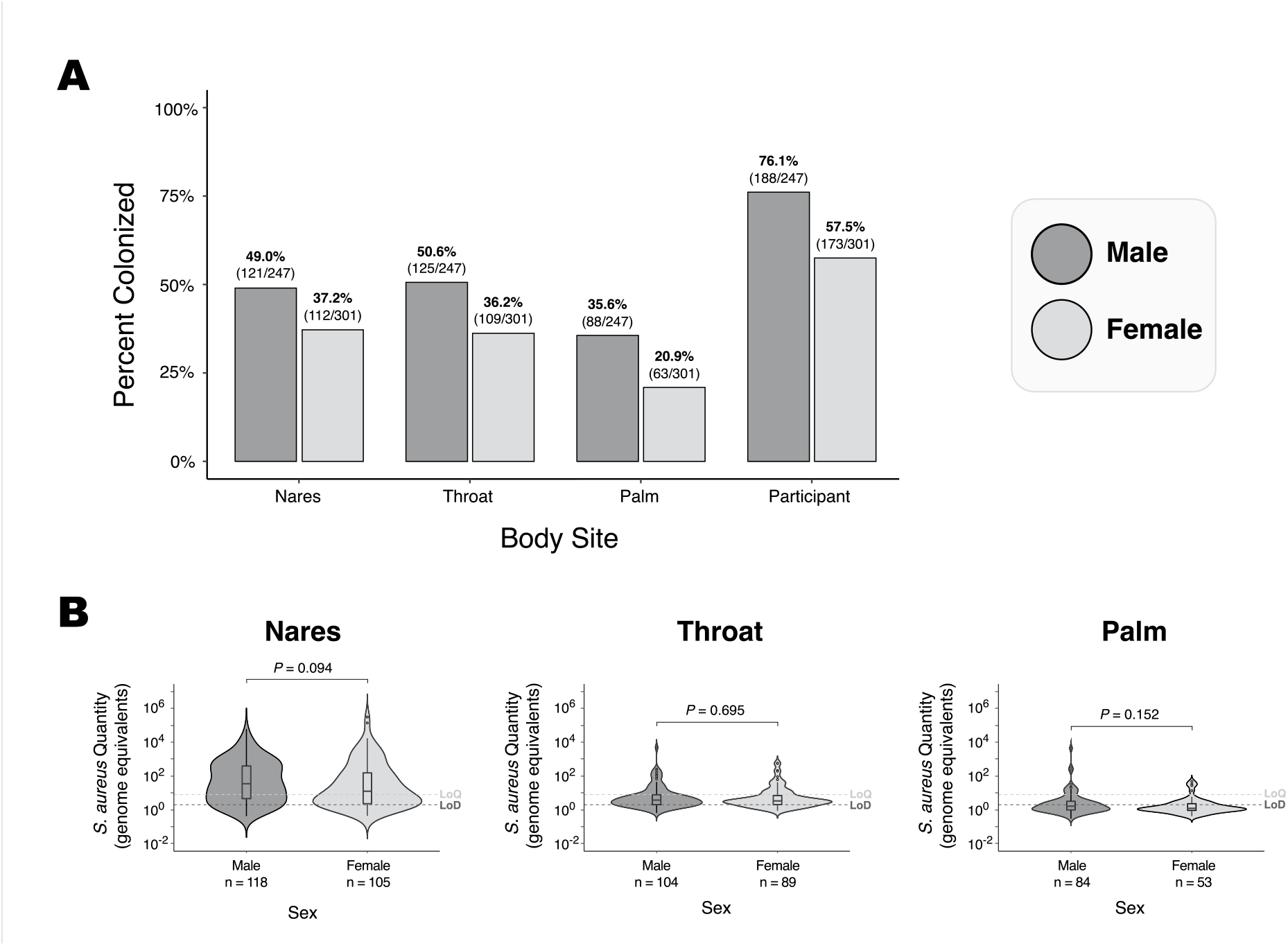
Colonization across sex and body site. A) Prevalence at different body sites for males and females shows a higher prevalence of *S. aureus* in males compared to females at all individual sites as well as on a participant level. Differences in prevalence between males and females were statistically significant according to the two-proportion z-tests (Nares: Z = 2.78, *P* = 0.006, Throat: Z = 3.39, *P* < 0.001, Palm: Z = 3.83, *P* < 0.001, Participant: Z = 4.58, *P* < 0.001). B) Differences in quantity between males and females were not statistically significant using the Mann-Whitney test. Quantity plots were made with a subset of the data used for the presence bar plots, as there are samples that were exclusively detected by culture, and hence have no associated quantity value.

*S. aureus* colonization across ethnicity was another area of interest considering findings suggesting a greater prevalence among non-Hispanic individuals (Kuehnert et al. 2006; Gorwitz et al. 2008). Our data revealed that *S. aureus* colonization was more prevalent for non-Hispanic participants compared to Hispanic participants at all body sites, although the difference was only statistically significant at the nares, throat, and participant level (Fig 5A). qPCR results showed significantly higher quantities for non-Hispanic individuals compared to Hispanic individuals at the nares (*P* = 0.028); However, differences at the palm and throat were not statistically significant (Fig 5B). Lastly, differences in *S. aureus* quantity and prevalence across participant age were also examined. Participants were split into three age classes (0-19, 20-49, and ≧50) to compare prevalence and quantity. *S. aureus* prevalence between age classes yielded no significant difference at any site, although the data generally shows higher prevalence among participants aged 20-49, and lower prevalence among participants aged 50 or greater (Fig 6A). Similarly, comparison of *S. aureus* quantity between age classes resulted in no significant difference at any body site (Fig 6B).

**Fig 5.**
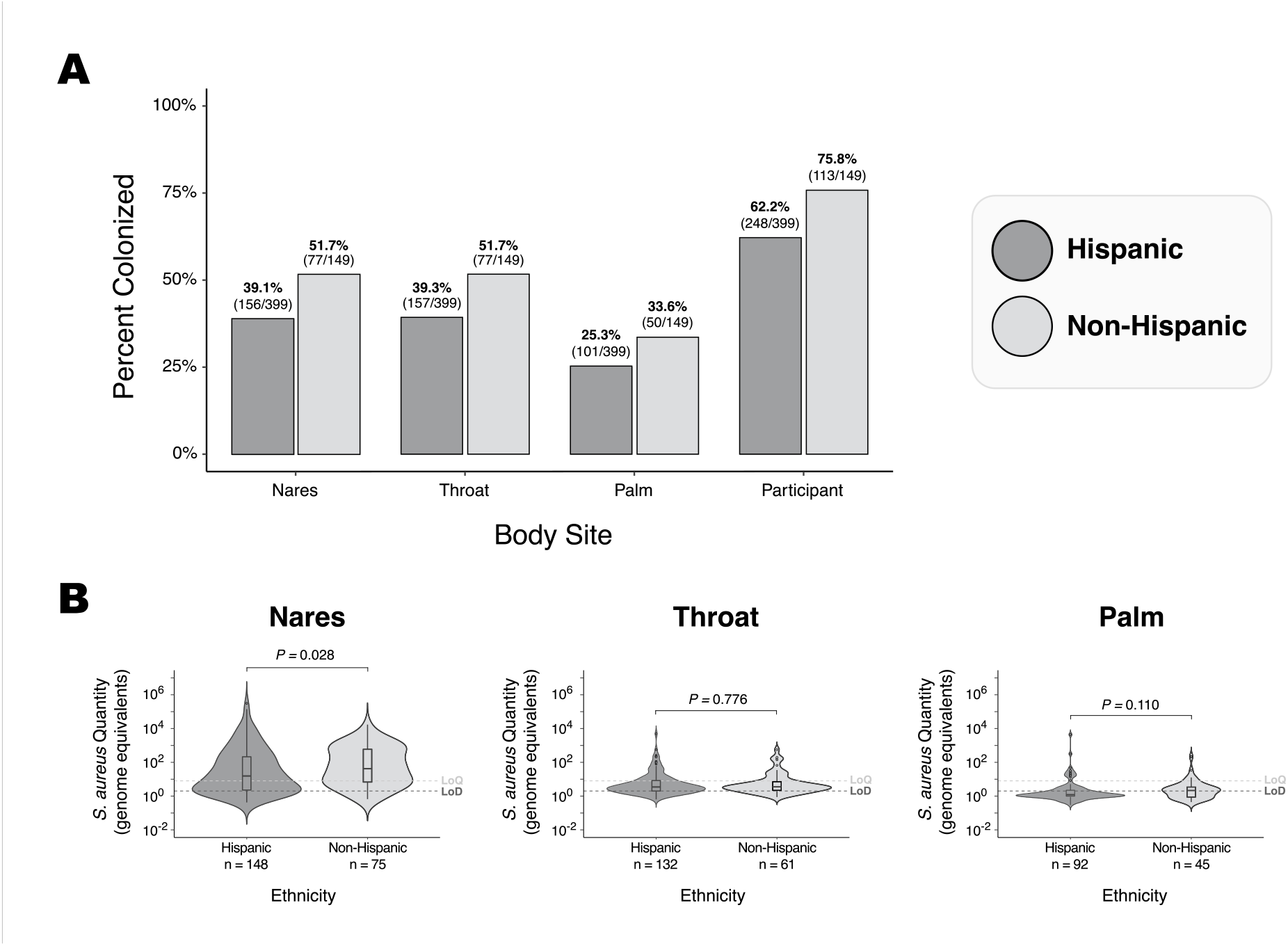
Colonization across ethnicity and body site. A) Non-Hispanic individuals had higher prevalence of *S. aureus* colonization compared to Hispanic individuals at the nares, throat, and participant level according to the two-proportion z-tests (Nares: Z = 2.65, *P* = 0.008, Throat: Z = 2.60, *P* = 0.009, Palm: Z = 1.92, *P* = 0.055, Participant: Z = 3.01, *P* = 0.003) (two-proportion z-test). Colonization was also higher in the palm for non-Hispanic individuals, but this difference was not statistically significant. B) *S. aureus* quantity was significantly higher for non-Hispanic individuals at the Nares body site.

**Fig 6.**
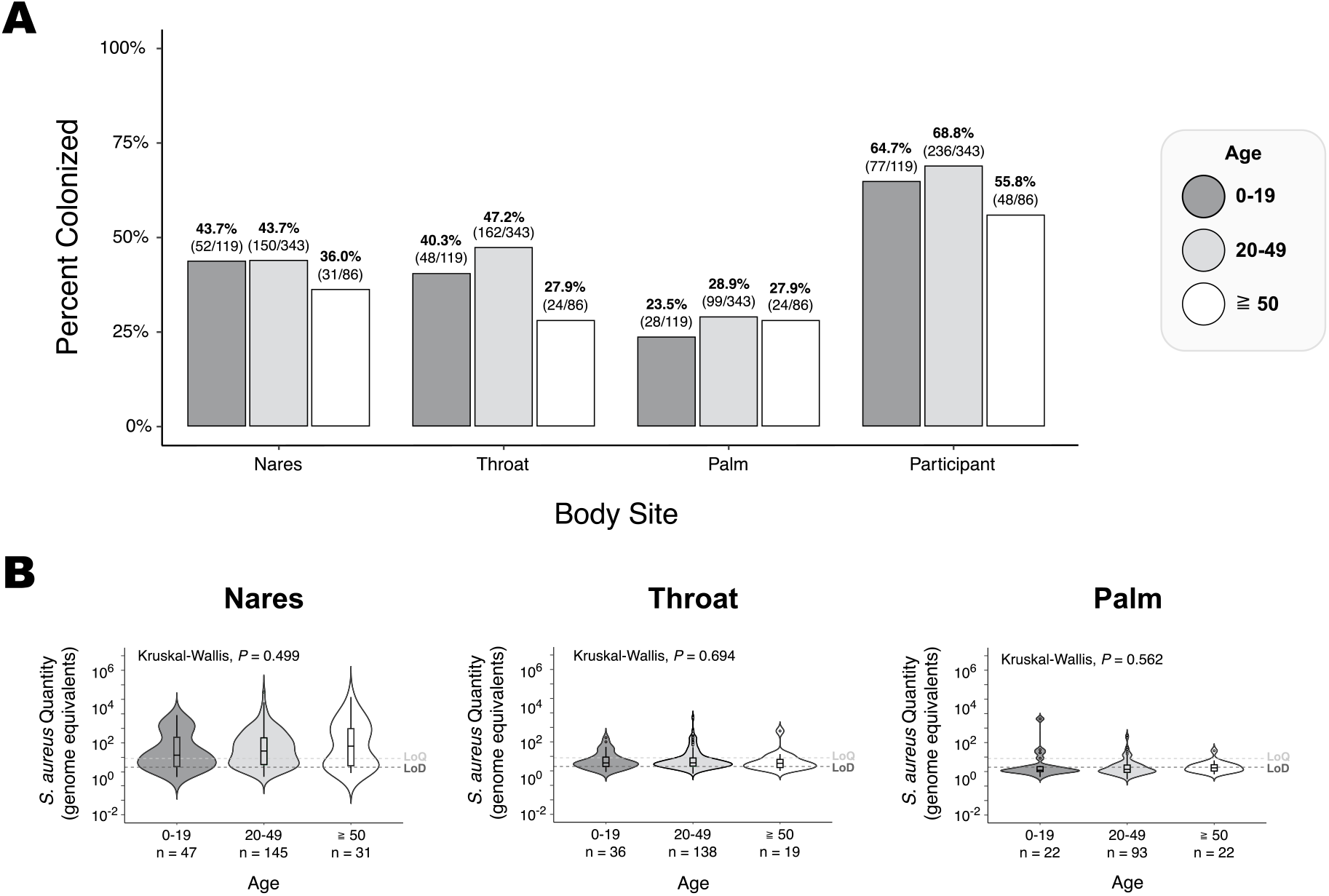
Prevalence and quantities at different body sites across three age classes. A) Prevalence patterns across different age groups suggests a lower prevalence in the oldest age group and a higher prevalence in the middle age group, but these differences are only significant at the throat and participant level between age groups 20-49 and ≧50 (Throat: *X*^*2*^ = 9.68, *P* = 0.002, Participant: *X*^*2*^ = 4.62, *P* = 0.032). All other comparisons between age groups 0-19 and 20-49 (Nares: *X*^*2*^ = 0, *P* = 1, Throat: *X*^*2*^ = 1.43, *P* = 0.232, Palm: *X*^*2*^ = 1.01, *P* = 0.316, Participant: *X*^*2*^ = 0.50, *P* = 0.478), 20-49 and ≧50 (Nares: *X*^*2*^ = 1.36, *P* = 0.243, Palm: *X*^*2*^ = 0, *P* = 0.967), as well as 0-19 and ≧50 (Nares: *X*^*2*^ = 0.92, *P* = 0.339, Throat: *X*^*2*^ = 2.86, *P* = 0.091, Palm: *X*^*2*^ = 0.30, *P* = 0.584, Participant: *X*^*2*^ = 1.31, *P* = 0.253), were not significantly different. B) Quantity of *S. aureus* at each body site does not differ according to age group.

### Cross-Site Influences

Sampling at multiple sites enabled us to conduct cross-site comparisons using data obtained from the SaQuant assay. Fig 7A shows the association between colonization at a given site and the presence and bacterial quantity of colonization at other sites. These comparisons reveal that a high *S. aureus* quantity in the nares is associated with concurrent colonization at other sites. Conversely, throat and palm quantity comparisons yielded no association with the number of sites colonized. Body site data also allowed us to study potential quantity associations between sites (Fig 7B). Comparisons showed no significant association between bacterial quantity for any pairwise comparison of body sites.

**Fig 7.**
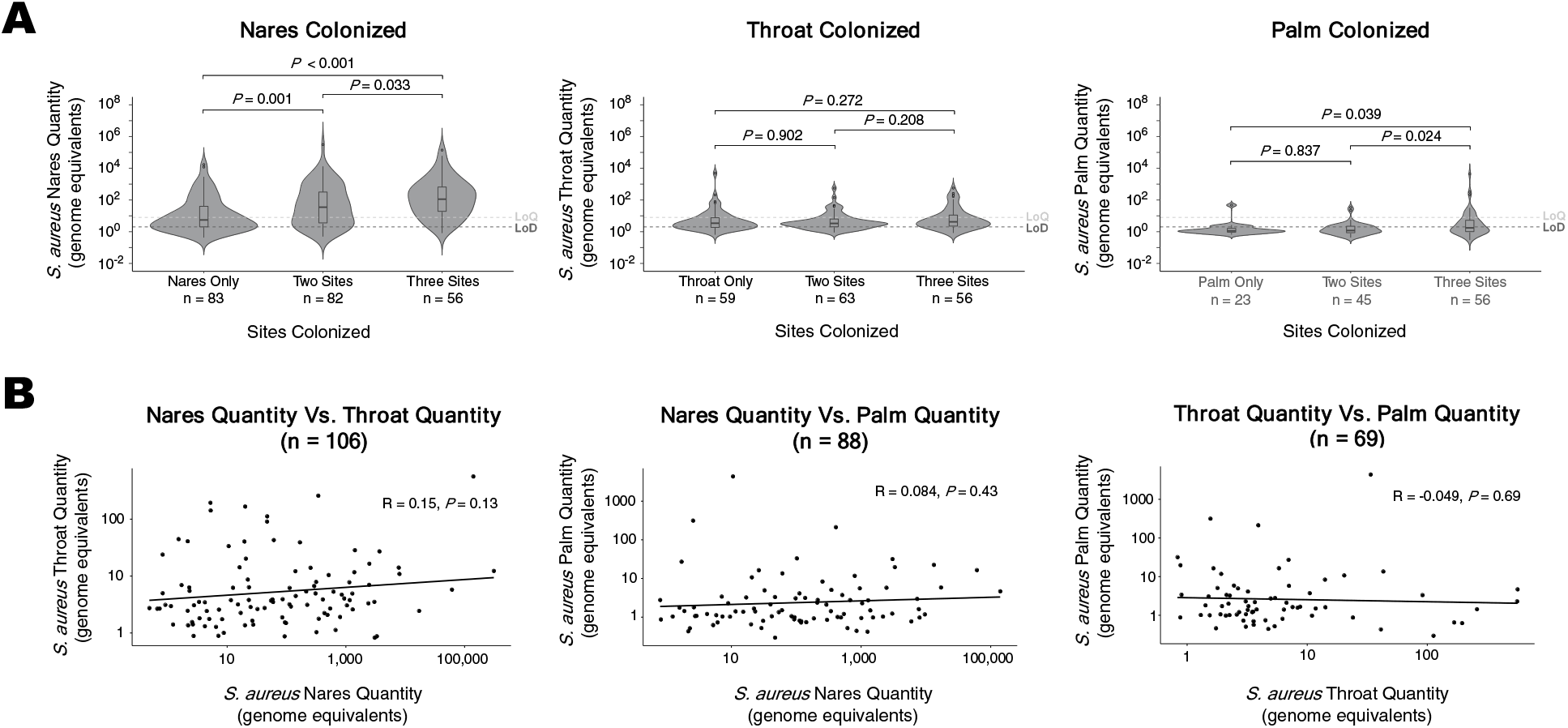
Association of colonization and bacterial quantity at one body site with colonization and bacterial quantity at other body sites. A) Quantity at each body site when different sites are also colonized. Quantity in nares was significantly higher when other sites were also colonized, however, B) quantities are not associated among sites.

## DISCUSSION

We combined culture and quantitative PCR-based methods on samples from three different body sites to characterize *S. aureus* colonization in a cohort of 548 participants in southwestern Arizona. Sampling from multiple body sites and combining culture with qPCR detection methods resulted in much higher sensitivity than sampling from any single site or culture alone. For example, combining all culture results (from the nares, throat, and palm) yielded a prevalence of 38% compared to 26.3% from nares cultures alone. Similarly combining culture and qPCR detection from nares samples yielded a prevalence of 42.5% compared to 26.3% and 40.7% for nares culture and nares qPCR prevalence (respectively) alone.

Overall, >65% of participants were colonized with *S. aureus*. Males and non-Hispanics were more likely to be colonized than females and Hispanics. Our current understanding of *S. aureus* carriage in the US population is largely based on data obtained in the 2001-2002 National Health and Nutrition Examination Survey (NHANES) which found an overall prevalence of ∼32%, with higher prevalence among males, but no clear difference between Hispanics and non-Hispanics (Kuehnert et al. 2006; Mainous et al. 2006; Gorwitz et al. 2008; Graham, Lin, and Larson 2006). Our data show a much higher overall prevalence and clear sex and ethnic-based differences. Slightly lower nasal prevalence, varying from 12.1% to 29.4%, were documented across 9 European countries, with higher prevalence in males and in younger age groups (den Heijer et al. 2013). However, both the US and European estimates, as well as those in other studies, were derived from culture-based detection from a single site, the anterior nares. In the present data, when also restricting positive results to culture-based detection from anterior nares swabs, overall prevalence (26.3%) is in line with previous estimates, suggesting that these previous studies are likely to have lacked sensitivity and underestimated prevalence.

Comparing prevalence across multiple body sites can improve insights into reservoirs and sources for spread to other body sites and other people. The anterior nares are most consistently identified as a site of colonization, providing evidence for the epidemiological importance of this body site. However, studies that consider both anterior nares and throat colonization show similar or even higher prevalence in the throat, suggesting that the throat should not be overlooked as a principal reservoir or dispersion site (Young et al. 2017; Erikstrup et al. 2019; Sollid et al. 2014; Hamdan-Partida, Sainz-Espuñes, and Bustos-Martínez 2010; Nilsson and Ripa 2006). Consistent with these studies, our data show that *S. aureus* was most prevalent in the nares (42.5%) and the throat (42.7%). Furthermore, 21.6% of individuals positive for *S. aureus* were colonized exclusively in the throat, which is slightly higher than the 19.9% colonized exclusively in the nares. *S. aureus* on the palm was least prevalent (27.6%), however 30 individuals (8.3%) were colonized exclusively on the palm, suggesting that the role of other body sites in maintenance and spread may be minor, but should not be ignored.

*S. aureus* quantity at any given site has direct implications on the likelihood of spread and transmission, and the likelihood of autoinfection (White 1961b; Ehrenkranz 1964), however we know almost nothing about quantitative aspects of community carriage. With one exception (Liu et al. 2015), the few studies that quantified *S. aureus* colonization relied on culture-based enumeration, but the expense and time-consuming nature of these methods have severely limited such studies. To address this deficit, we utilized SaQuant, a recently developed assay for the detection and quantification of *S. aureus* (Wood et al. 2021). A small number of other PCR assays are available for *S. aureus*; however, they were 1) not validated for quantification, 2) used limited *in silico* data (compared to what is available today), 3) utilized proprietary DNA signatures that cannot be readily assessed, 4) include assay metrics that were not peer reviewed, or 5) have lower sensitivity/specificity (Wood et al. 2021). By employing this quantitative assay alongside culture-based detection, we not only show that culture is less sensitive than SaQuant, but also that this failure to detect positive samples was mostly due to low quantities of *S. aureus*. Our use of SaQuant for quantitative assessment of *S. aureus* colonization reveals some interesting characteristics of carriage in the anterior nares compared to the throat and palm. For example, quantity in the anterior nares is highly variable, with values ranging from less than a single genome equivalent per 1μL to over 310,386. The average quantity in the nares was significantly greater than the throat, which in turn, was greater than the palm. Unlike the other body sites, the average quantities in the nares were greater in males (compared to females) and non-Hispanics (compared to Hispanics), although the sex-based difference was not statistically significant. Quantities in the nares were also greater when other sites were colonized, however quantities in those other sites were not correlated with nares quantity. These characteristics of nares colonization (the potentially massive quantities that may be found in the nares, the correlation of quantities in the nares with sex, ethnicity, and colonization at other body sites) were not observed for the throat and suggest that the epidemiological role of nares colonization may be significantly distinct from other sites.

Our work has some important limitations. Firstly, culture-based detection methods for *S. aureus* are varied and our detection sensitivity may have been improved by broth enrichment for example. Similarly, sensitivity for qPCR-based detection would have been increased by testing replicates to reduce the stochasticity associated with detecting low target quantities. As a result, our prevalence values for both culture and qPCR are undoubtedly underestimated. Secondly, our sampling was cross-sectional, and thus does not provide any insights into carriage over time or temporal relationships between carriage across sites. Lastly, unlike other studies (Gorwitz et al. 2008; Graham, Lin, and Larson 2006; Erikstrup et al. 2019; Holtfreter et al. 2016), we did not observe any statistically significant age-based differences in colonization, however this may be due to small sample sizes in different age groups or underestimation of detection in these other studies.

## Conclusions

Effective disease control and prevention is dependent on identifying and understanding pathogen reservoirs. A major contribution of our work is that previous efforts are likely to have lacked sensitivity for detection of *S. aureus* and therefore underestimated carriage. There exists a large body of literature describing prevalence of *S. aureus*, but it is difficult to compare results across studies due to different collection, storage, and culture methodologies, different sampling timelines, and different subject demographics. Our study allows for some comparisons to previous work as we sampled from commonly targeted body sites and used a culture-based detection method in addition to molecular detection. While our culture-based prevalence results are consistent with expectations based on previous studies, our qPCR detection results are much higher than has been documented in a general population (J. Kluytmans, van Belkum, and Verbrugh 1997; den Heijer et al. 2013). It is certainly possible that our population is anomalous for some unknown reason, however our data provide evidence that our high overall prevalence estimates are explained by higher detection sensitivity (assessing 3 body sites and utilizing direct culture in combination with qPCR). The failure of culture-based methods to capture a substantial portion of carriers has been previously suggested (Liu et al. 2015) and the gains in sensitivity by using PCR and sampling multiple body sites for the detection of MRSA has been demonstrated (Senn et al. 2012). Taken together, previous prevalence studies that sampled a single body site or relied on culture-based detection may have significantly underestimated carriage. These studies however form the foundation of our current understanding of community carriage, longitudinal carriage within individuals, carriage at different body sites, assessments of infection risk, and infection mitigation practices. Pervasive underestimation of carriage and insufficient practices for identifying carriage may, at least partially, explain why infection control progress seems to have stalled.

Few studies have investigated the epidemiological importance of carriage quantity and this work helps fill this knowledge gap using a new high-throughput quantitative tool. These studies suggest that a high quantity of *S. aureus* is more likely to be found in the nares compared to the axillae and groin (Rohr et al. 2004), in persistent carriers (White 1961a), and males (Liu et al. 2015), leads to higher environmental contamination (White 1961b), colonization at other body sites (Mermel et al. 2011), transmission to others (Ehrenkranz 1964) and ultimately, a higher risk of infection (Calia et al. 1969; White 1963). We are aware of only a single study of carriage quantity in a non-clinical population (Liu et al. 2015). While providing profound insights, these few studies do not provide a comprehensive understanding of the epidemiological role of carriage quantity and the general lack of high-throughput tools for quantifying *S. aureus* in a sample has limited research efforts. We used SaQuant, a new qPCR assay with high sensitivity and specificity to more fully characterize carriage by documenting how carriage quantity varies across body sites, sex, and ethnicity in a large non-clinical cohort in southwestern Arizona.

Efforts to mitigate *S. aureus* infections have been met with diminishing returns. Progress has certainly been made in reducing infections in healthcare environments, but perhaps the relatively high remnant infection rates are due to the persistence of small quantities in various body sites that go undetected despite screening and decolonization efforts. Our work demonstrates the importance of sensitive and quantitative detection in understanding carriage. Longitudinal sampling over a greater diversity of body sites and populations will be critical for more comprehensive understanding of colonization and will provide further guidance towards more effective prevention of *S. aureus* infections.

### Ethics approval and consent to participate

Samples tested in this work were collected as a part of project 1116783 approved by the Northern Arizona University Institutional Review Board. Verbal consent or assent was obtained to maintain anonymity of participants.

## Data Availability

All data produced in the present work are contained in the manuscript

## Acknowledgements

This work would not have been possible without the support from the Regional Center for Border Health in Somerton, AZ. We are grateful to the many field surveyors from NAU and the Regional Center for Border Health who recruited, sampled, and collected data from participants. This work was supported by the NAU Southwest Health Equity Research Collaborative, which is funded by the National Institute on Minority Health and Health Disparities at the National Institutes of Health (U54MD012388). Funding was also received from the National Institute of Allergy and Infectious Diseases at the National Institutes of Health (R15AI156771). The funders had no role in study design, data collection and analysis, decision to publish, or preparation of the manuscript.

